# Simulated Reasoning and Self-Verification for Psychiatric Diagnosis in Generalist Large Language Models: Comparative Evaluation

**DOI:** 10.1101/2025.09.05.25335196

**Authors:** Karthik V Sarma, Kaitlin E Hanss, Andrew J M Halls, Daniel F Becker, Anne L Glowinski, Andrew Krystal

## Abstract

**Background:** Large language models (LLMs), and, more recently, large reasoning models (LRMs) have rapidly garnered significant interest for application in psychiatry and behavioral health. However, recent studies have identified significant shortcomings and potential risks in the performance of LLM-based systems, complicating their application to psychiatric diagnosis. Two promising approaches to addressing these challenges and improving the efficacy of these models are simulated reasoning (SR) and self-verification (SV), in which additional “reasoning tokens” are used to guide model output, either during or after inference.

**Objectives:** We aimed to explore how the use of SR (via LRMs) and SV (via supplemental prompting) affect the psychiatric diagnostic performance of LLMs.

**Methods:** 106 case vignettes and associated diagnoses were extracted from the DSM-5-TR Clinical Cases book, with permission. Both an LLM and LRM model were selected from the latest available model generation for each of the two vendors studied (OpenAI and Google). Two inference approaches were developed, a Basic approach that directly prompted models to provide diagnoses, and a SV approach that augmented the Basic approach with additional prompts. All case vignettes were processed by the two LLMs, two LRMs, and two inference approaches, and diagnostic performance was evaluated using the sensitivity and positive predictive value (PPV). Binomial generalized linear mixed models were used to test for significant differences between the model vendors (OpenAI, Google), type (LLM, LRM), and addition of an SV prompt.

**Results:** All vignettes were successfully processed by each model and inference approach. Sensitivity ranged from 0.732 to 0.817, and PPV ranged from 0.534 to 0.779. The best overall performance was found in the *o3-pro* LRM using SV, with a sensitivity of 0.782 and a PPV of 0.779. No statistically significant fixed effects were found for sensitivity. For PPV, a statistically significant effect was found for prompt type (SV, p=0.007), model type (LRM, p=0.009). No significant interaction effects were identified.

**Conclusions:** We found that both SR and SV yielded statistically significant improvements in the PPV, without significant differences in the sensitivity. The addition of the manually specified SV prompt improved the PPV even when simulated reasoning was used. This suggests that future efforts to apply language models in behavioral health could benefit from manually crafted reasoning prompts and automated SR.

## Introduction

Large language models (LLMs) have rapidly garnered significant interest for application in psychiatry and behavioral health. LLMs, a type of deep artificial intelligence (AI) system, are trained as “next token” (i.e., next word) predictors on very large-scale unsupervised text corpora, with estimates in the trillions of tokens. [1] These models have demonstrated unexpected and remarkable emergent capabilities in the areas of information retrieval and processing and problem-solving, suggestive of a capability to engage with the semantic content of unstructured text not possible with previous technologies.

These emergent capabilities have enabled the demonstration of both clinical and consumer-facing LLM-based systems with a broad set of applications across fields of medicine. In behavioral health, projects have demonstrated their use in automated diagnostic and therapeutic interactive agents (i.e., chatbots) [2], question-answering [3], and diagnostic reasoning. [4] However, recent studies have also identified significant shortcomings and potential risks in the performance of LLM-based systems, including hallucination, bias, sycophancy, and the generation of associated misaligned and dangerous content for users experiencing mental health challenges. [5–8] These challenges complicate the application of large language models to psychiatric diagnosis, especially in the context of the variability in interrater reliability of DSM diagnoses. [9,10] Though some published work has noted the efficacy of structured prompt-based decision processes [4], other works have found that structured behavioral health prompts, such as psychometric scales, may not work effectively with generalist models. [11]

Two promising approaches to addressing these challenges and improving the efficacy of these models are simulated reasoning (SR) and self-verification (SV). In these methods, a structured approach is used to generate additional tokens (known as *reasoning tokens*), either generated by the model or provided by the system designer, that increase the probability of correct response. In SR, tokens are generated to create a reasoning “chain of thought” that leads to the model’s final response. In SV, once a preliminary response is generated by the model, reasoning tokens are added to lead the model to re-evaluate the correctness of this answer before generating a final result. LLMs that automatically incorporate the use of reasoning tokens are often known as large reasoning models (LRMs), and the major vendors of LLMs have now produced LRM versions of their latest-generation tools (i.e., OpenAI o3, Google Gemini 2.5 Pro). Some recent works, however, have suggested that the use of reasoning tokens in LRM may not lead to reasoning-like processes [12–14] in some settings. For example, in one evaluation by a major vendor, researchers found that reasoning tokens output by an LRM frequently did not represent actual reasoning processes, as evidenced by their failure to note direct hints that were provided in initial prompts. [13] In contrast, a large vendor recently stated that they plan to automatically move conversations that contain acute distress into reasoning models for “more helpful and beneficial responses” in these circumstances. [15]

Though the exploration of LLM-based tools for clinical application continues to progress rapidly, few studies are available to guide the choice of model or reasoning approach specifically for behavioral health, or to provide estimation of how these models will perform in this setting. There is an expanding body of evidence on the use of these tools across other fields of medicine, with varied findings. In one recent paper, a selection of LLMs and LRMs were tasked with clinical reasoning using vignettes across domains of medicine; this work found high accuracy on final diagnosis but poor performance in other reasoning stages [16]; another study similarly attempted to assess diagnostic reasoning across domains of medicine using script concordance testing and found sub-clinician reasoning performance [17]. Neither study specifically investigated the differential impact of simulated reasoning on outcomes. Several efforts have worked towards developing benchmarks to enable serial evaluation of performance, without a focus on behavioral health [18,19].

In prior work, we demonstrated that fixed prompt decision trees improve LLM psychiatric diagnostic performance [4] in a vignette dataset, without assessing LRMs. A recent unpublished preprint similarly used a vignette dataset to assess diagnostic performance, making use of state-of-the-art LRMs and finding moderate to high diagnostic accuracy, without specifically investigating the differential impact of methods like SR or SV. Another study examined the use of LLMs with real-world EHR data, finding promising but variable diagnostic performance, without investigating methodological factors. [20]

To address the evidence gap in understanding the impact of differing reasoning methods for behavioral health, we aim here to evaluate whether the use of SR (i.e., LRMs) and/or SV (i.e., standardized post-processing) enhances LLM performance on a vignette-based diagnostic task.

## Methods

### Standardized Task and Dataset

We extracted 106 case vignettes written by 133 discussants across 19 primary DSM-5-TR diagnostic categories from the DSM-5-TR Clinical Cases book. Each vignette is a multi-paragraph description of a particular case; the vignettes are written for educational purposes; the author’s intended final diagnoses are also provided separately from the vignette. The standardized task was to generate diagnoses from the case vignette, with the author-designated diagnoses used as the gold-standard comparison.

#### Memorization Analysis

To assess the potential impact of the models’ possible prior knowledge of these vignettes, we conducted a two-part memorization analysis. For each of the 106 vignettes, we constructed two truncation points: an early truncation (after sentence 3) and a midpoint truncation (at the halfway point of the vignette by sentence count). In separate queries for each truncation point, the model was provided with vignette text up to that point and prompted to provide the following exact sentences or, if unable, to write a plausible continuation. The exact prompt was as follows:

The following is the beginning of a passage of text. Please continue the passage by writing exactly the next 2-3 sentences as they appear in the original source. If you do not recognize the text, write a plausible continuation. <TEXT>

This was performed independently for all of the LLMs and LRMs used in the study. Three similarity metrics were computed for each model-generated continuation by comparison to the vignette: ROUGE-L [21], BLEU-4 [22], and exact sentence match.

### Model Selection and Configuration

For this study, we selected one LLM and one LRM from two major vendors from whom an enterprise-grade LLM application programming interface was available, and which had agreed terms of use that were 1) compatible with the IP rights of the study researchers and rightsholders associated with the study data, and 2) prohibited the use of study data for model training by vendors. The vendors and models used for this study were: OpenAI (LLM: *gpt-4*.*1-2025-04-14*, LRM: *o3-pro-2025-06-10*), and Google (LLM: *Gemini 2*.*5 Flash*, LRM: *Gemini 2*.*5 Pro*). The LLMs were used at their default settings, with an output token limit of 2000. LRMs were provided with 16,000 reasoning tokens, and OpenAI models were also set to medium reasoning effort (Google models do not provide this parameter).

Reasoning was disabled for both LLMs. Safety settings were disabled or set to the minimum level of filtering. Models were constrained to respond in a specified machine-parsable output format (JavaScript Object Notation, or JSON) either through the use of application programming interface (API) flags or by re-prompting until the output was in valid JSON. All prompts and re-prompts were independent without inclusion of prior input, output, or logs.

### Inference Approaches

We compared two inference approaches previously developed by the authors [4] using each of the available models. In the first approach (the “Basic” approach), the model was directly prompted to assign diagnoses to the vignette using a standardized prompt (Table 1). In the second approach (the “SV” approach), diagnoses generated by the Basic approach were evaluated using a sequential pairwise elimination procedure. Each pair of candidate diagnoses was presented to the model using a standardized prompt asking whether both diagnoses were necessary or whether one was better explained by the other (Table 1); each of these prompts was done independently without including context from any other prompt. If both were retained, the procedure continued; if only one was selected, the other was removed from further evaluation. Because this procedure requires at least two candidates, vignettes for which the Basic approach produced a single diagnosis were not modified by SV, and if a candidate diagnosis list was reduced to only one candidate, the procedure was terminated. This approach serves as a templated unidirectional filter that can only narrow the list of diagnoses.

**Table 1.** Prompt Templates for Inference Approaches.

|  |
| --- |
| <u>Basic Prompt</u><br>I am going to give you an academic psychiatry clinical case that describes a patient with one or more psychiatric DSM-5-TR diagnoses, and you are going to answer questions about that case that I provide.<br>The clinical case is as follows: <X><br>Please provide me with a list of DSM-5-TR diagnoses, without specifiers or modifiers, that you believe apply to this patient based solely on the clinical case. Please format them as a JSON list titled "diagnoses" with one diagnosis per entry. Do not include any other text in your response. Do not include any incorrect, inappropriate, or candidate diagnoses. For example, if the diagnoses are "Insomnia Disorder" and "Bipolar I Disorder", you would reply with: {"diagnoses": ["Insomnia Disorder", "Bipolar I Disorder"]} |
| <u>Self-Verification Prompt</u><br>The following two DSM-5-TR diagnoses are candidate diagnoses for this patient: <X> and <Y>. We are interested in deciding if both diagnoses are necessary for the patient, or if one of these two diagnoses is better explained by the other. Please respond with a list of which of these two diagnoses are necessary for the patient in JSON format. For example, if the candidate diagnoses are 'major depressive disorder' and 'adjustment disorder', respond with ["major depressive disorder", "adjustment disorder"] if both diagnoses are necessary, or with either ["major depressive disorder"] or ["adjustment disorder"] if one diagnosis is better than the other. |

### Scoring and Analysis

#### Diagnosis Matching, Simplification, Scoring, and Metrics

To facilitate ease of comparison of model-generated and author-designated diagnoses, a previously developed semi-automated standardized diagnosis matching and simplification system was applied to all diagnoses. In this system, all specifiers and modifiers were removed, neurocognitive disorders were collapsed into either a catch-all diagnosis for delirium or a catch-all diagnosis for major or minor neurocognitive disorders, and then diagnoses were systematically matched to DSM-5-TR diagnoses, Z codes, or other categories. See Multimedia Appendix 1 for full detail on this matching system. A model-generated diagnosis was scored as a true positive (TP) if it matched an author-designated diagnosis, or as a false positive (FP) otherwise; author-designated diagnoses without a matching model prediction were scored as false negatives (FN). Per-vignette sensitivity was calculated as TP/(TP+FN) and PPV as TP/(TP+FP).

#### Statistical Analysis

Sensitivity and PPV were averaged across vignettes for reporting, and 95% confidence intervals were obtained by bootstrap resampling with replacement (10,000 iterations). During bootstrap computation shared indices were used across all experiments to ensure consistency. Macro-averages were computed for reporting, giving equal weight to each vignette, to align with statistical modeling. Micro-averages were also computed to assess for consistency.

For statistical comparison, we fit count-level (i.e., TP, FP, FN) binomial generalized linear mixed models (GLMMs) to sensitivity and PPV, using a logit link and random intercept in *R*, to examine the fixed effects of model vendor (OpenAI, Google), type (LLM, LRM) and use of the SV prompt; two- and three-way interactions were also examined. A threshold of *p* < 0.05 was used to evaluate for significant fixed effects.

## Results

All 106 vignettes were processed by the four models, without any parsing errors or context window length overruns that would have required re-prompting. The memorization analysis was reassuring against memorization; no model produced any exact sentence match and ROUGE-L and BLEU-4 scores demonstrated minimal overlap (Table 2). There were between 0 and 5 author-designated diagnoses per vignette distributed as: 0 (0.9%), 1 (58.5%), 2 (30.2%), 3 (7.5%), 4 (1.9%), and 5 (0.9%); the average number per vignette was 1.54. Two cases were not processed by the Google models due to a content filter block; both cases were treated as if they had zero predicted diagnoses.

**Table 2.** Memorization Analysis by Model.

| Vendor | Model Type | ROUGE-L mean ± SD | BLEU-4 mean ± SD |
| --- | --- | --- | --- |
| OpenAI | LLM | 0.154 ± 0.040 | 0.013 ± 0.012 |
| Google | LLM | 0.153 ± 0.047 | 0.013 ± 0.019 |
| OpenAI | LRM | 0.106 ± 0.047 | 0.007 ± 0.011 |
| Google | LRM | 0.153 ± 0.044 | 0.013 ± 0.014 |

LLM = Large Language Model; LRM = Large Reasoning Model

Simplification and matching were completed and all model-predicted diagnoses were matched to a DSM-5-TR diagnosis, Z code, or a non-psychiatric diagnosis. There was one case (7.2.1) with zero author-designated diagnoses (i.e., no diagnosis was applicable); in this case, the sensitivity was undefined and was set as 0 for all models as all models incorrectly generated at least one false positive diagnosis for this case. There were no cases in which the PPV was undefined (i.e., the model generated no diagnoses). Sensitivity ranged from 0.732 to 0.817, PPV ranged from 0.534 to 0.779. Macro-averaged values for sensitivity and PPV for each experiment are available in Table 3; micro-averaged values were computed and found to provide qualitatively similar results. Total counts across each category for each experiment are available in Table 4. The Basic approach generated an average of 1.73 to 2.42 diagnoses per vignette across all four models, which SV reduced to 1.57 to 1.89. Per-vignette performance is depicted in Figure 1 and individual vignette-level results are available in Multimedia Appendix 2.

**Table 3.** LLM Diagnostic Performance Metrics by Experiment.

| Vendor | Model Type | Inference Approach | Sensitivity | PPV |
| --- | --- | --- | --- | --- |
| Google | LLM | Basic | 0.77 [0.68-0.84] | 0.53 [0.47-0.60] |
|  |  | SV | 0.75 [0.68-0.82] | 0.63 [0.56-0.70] |
|  | LRM | Basic | 0.79 [0.72-0.86] | 0.63 [0.56-0.69] |
|  |  | SV | 0.77 [0.70-0.84] | 0.67 [0.60-0.74] |
| OpenAI | LLM | Basic | 0.75 [0.67-0.82] | 0.57 [0.50-0.64] |
|  |  | SV | 0.73 [0.66-0.80] | 0.64 [0.57-0.71] |
|  | LRM | Basic | 0.82 [0.75-0.88] | 0.76 [0.69-0.83] |
|  |  | SV | 0.78 [0.71-0.85] | 0.78 [0.71-0.85] |

LLM = Large Language Model; LRM = Large Reasoning Model; SV = Self-Verification; PPV = Positive Predictive Value

**Table 4.** Diagnosis Statistics by Experiment.

| Vendor | Model Type | Inference Approach | Author Dx | Model Dx | TP | FP | FN |
| --- | --- | --- | --- | --- | --- | --- | --- |
| Google | LLM | Basic | 163 | 257 | 124 | 133 | 39 |
|  |  | SV | 163 | 200 | 121 | 79 | 42 |
|  | LRM | Basic | 163 | 213 | 127 | 86 | 36 |
|  |  | SV | 163 | 188 | 121 | 67 | 42 |
| OpenAI | LLM | Basic | 163 | 233 | 125 | 108 | 38 |
|  |  | SV | 163 | 192 | 120 | 72 | 43 |
|  | LRM | Basic | 163 | 183 | 135 | 48 | 28 |
|  |  | SV | 163 | 166 | 126 | 40 | 37 |

LLM = Large Language Model; LRM = Large Reasoning Model; SV = Self-Verification; Author Dx = Author-Designated Diagnoses; TP = True Positive; FP = False Positive; FN = False Negative.

**Figure 1.**
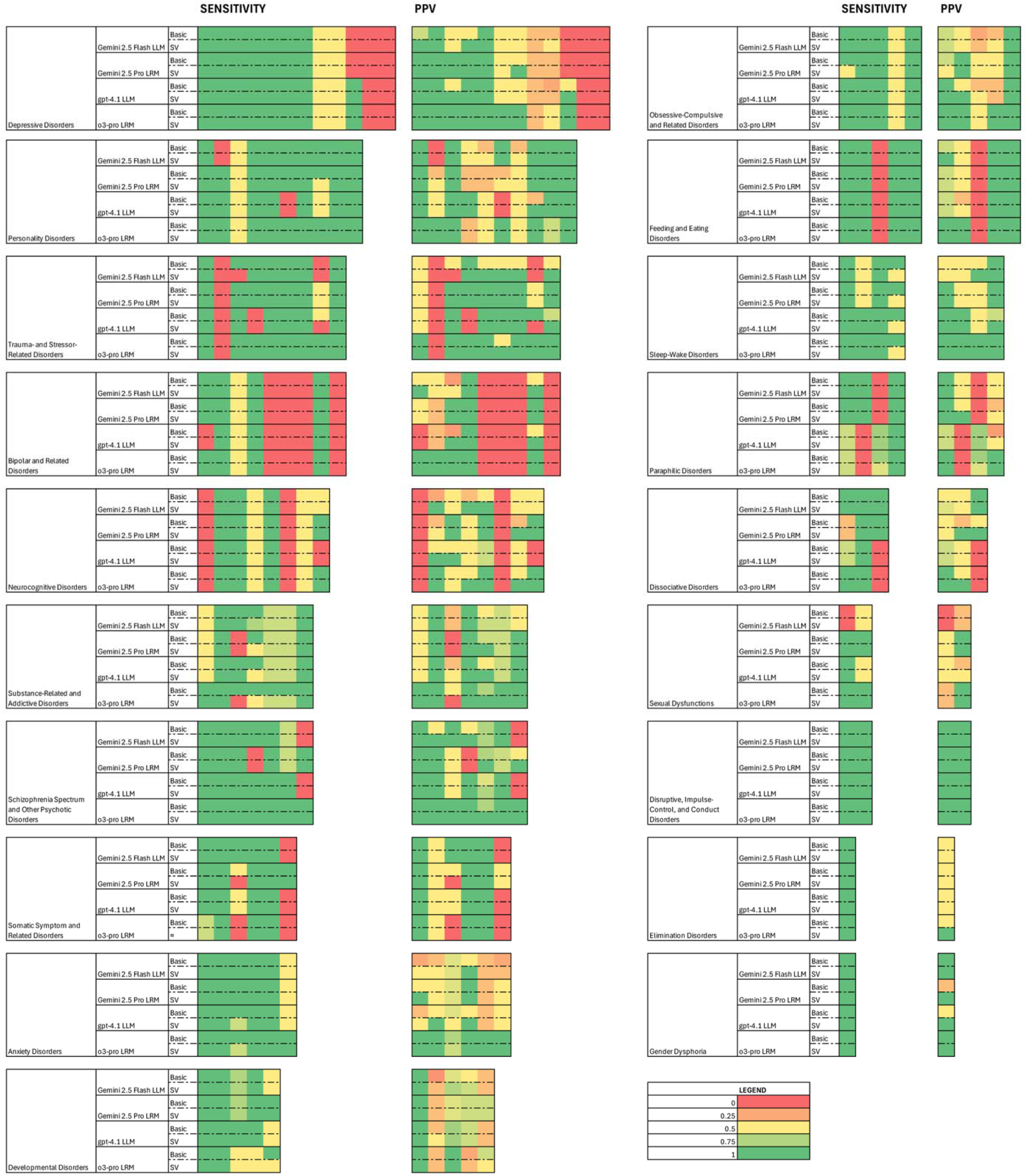
Per-case results of evaluation of diagnostic performance, by experiment and diagnostic category. This color map displays the per-case sensitivity and positive predictive value (PPV) for diagnosis for every vignette, grouped by DSM-5-TR category. Each row consists of experimental results in one specific category for one specific experiment (i.e., combination of model and inference approach). A legend is displayed for the color map. Each column within each metric represents a single specific case vignette.

Binomial GLMMs were successfully fit. The intraclass correlation (ICC) was 0.77 for sensitivity and 0.49 for PPV. No statistically significant fixed effects were found for sensitivity. For PPV, a statistically significant effect was found for prompt type (SV, p=0.007), model type (LRM, p = 0.009). No statistically significant interactions were identified.

## Discussion

In this work, we sought to evaluate the impact of SR and SV on the psychiatric diagnostic performance of language models from two different vendors.

### Principal Findings

We successfully processed 106 psychiatric case vignettes using the selected approaches and found that both simulated reasoning and self-verification yielded statistically significant improvements in the PPV, compared to the Basic approach alone, without significant differences in the sensitivity. The best overall performance of models was found in the *o3-pro* LRM with the additional SV prompt, yielding a sensitivity of 0.782 and a PPV of 0.779. The ICC findings may suggest that much of the variance found in our experiments was explained by the intrinsic difficulty of the individual vignettes, a finding also clear in the per-vignette results depicted in Figure 1; however, for PPV, substantial variance is likely attributable to differences in approach (since, for example, the SV prompt acts as a unidirectional filter, only removing diagnoses).

Interestingly, we found no statistically significant interaction effect between SR and SV, suggesting that the use of both yielded no additional benefit over any one alone. This finding may, however, be due to a lack of sufficient power from our study to detect these differences and future evaluation is required. Thus, the addition of explicit reasoning suggestions in prompts (such as that in the SV prompt) could continue to be helpful even when using automated simulating reasoning systems. Similarly, we found no significant impact of the use of either approach on sensitivity; this is unsurprising for SV given that this approach can only remove diagnoses, but was surprising for SR which theoretically could use reasoning tokens to expand diagnostic range.

### Limitations

First, the semi-automated simplification and matching process we used was intended to give models the best opportunity to be scored as correct, and this may have the impact of overestimating the performance of the models. Second, the vignettes and diagnoses used for our experiment may be in the training set of these models (though they have not been licensed for such use); our test results are reassuring against memorization and the vignettes are explicitly not licensed for this use, but it is still possible that training exposure may have biased our evaluation towards better performance. Even in this case, however, we believe that the relative findings between methods in our study would be valuable. Third, since the vignettes were written for teaching, the prevalence of diagnoses is higher in the dataset than in the general population and more comprehensive information is provided in the vignettes than is generally available in the clinical setting. This may serve to inflate the performance metrics that we found and our results are not intended to suggest clinical utility at this stage. Additionally, the amplified illness prevalence in a vignette-based educational dataset enables methodologic exploration, but findings may not generalize to low-prevalence real-world populations (such as self-diagnosing internet users). Fourth, not all diagnostic categories are equally or proportionally covered by our dataset, and our study was not powered to detect inter-category differences. Additionally, because we specifically collapsed neurocognitive disorders into two diagnoses (Multimedia Appendix 1), performance on these cases may be artificially inflated by masking potential challenges in differentiating between specific neurocognitive pathologies.

### Future Directions

Future efforts could make use of larger datasets, potentially including enhanced coverage of each diagnostic category and making use of either novel vignettes or real-world clinical data, considering full differential diagnosis, and comparison to human performance. This would enhance the power of future studies and minimize the risk of training bias. Additionally, future studies could examine the impact of model selection and prompt optimization to better elucidate how best to configure language models for psychiatric use, including the potential blending of SR and SV-like approaches, such as the use of pre-templated prompts that call for generating additional reasoning prompts.

## Conclusion

In this study, we found that both simulated reasoning and self-verification improved the positive predictive value of LLM-based psychiatric diagnosis from case vignettes, without significantly affecting sensitivity. The impact of these two approaches suggests that manually crafted verification prompts can continue to provide value even when automated reasoning capabilities are available. Future work should validate these findings using real-world clinical data in order to inform clinical system design.

## Supporting information

Multimedia Appendix 1

Multimedia Appendix 2

## Acknowledgements

The authors acknowledge the mentorship and scientific vision provided by Dr. Atul J Butte, MD PhD, director of the UCSF Bakar Computational Health Sciences Institute, who passed away in June 2025. We thank the UCSF AI Tiger Team, UCSF Academic Research Services, UCSF Research Information Technology, and the UCSF Chancellor’s Task Force for Generative AI for their support in developing LLM resources used for this project. Generative AI was used as described in the methods section for the execution of this research. It was not used in the production of the initial draft manuscript, but was used to assist in the development of revisions, including by reviewing and suggesting edits to address reviewer comments and generating candidate language for inclusion in the manuscript. All content was manually reviewed by a human author and the authors take responsibility for all content in this manuscript. The authors appreciated the opportunity to submit an earlier component of this work to the 2026 ACNP Annual Meeting. The contents of this manuscript are solely the responsibility of the authors and do not necessarily represent the official views of the NIH, APA, UCSF, or any other organization.

## Data Availability

The dataset used for this study was extracted from the DSM-5-TR Clinical Cases textbook under license from the American Psychiatric Association (as follows), and cannot be reproduced by the authors of this study:

This research was made possible through the use of content belonging to the American Psychiatric Association; express permission was obtained from the American Psychiatric Association for the use of such content (DSM-5-TR Clinical Cases. Copyright © 2023. American Psychiatric Association. All Rights Reserved, including rights for text and data mining (TDM), Artificial Intelligence (AI) training, and similar technologies).

## Authors’ Contributions

Karthik V Sarma: conceptualization, methodology, software, validation, formal analysis, investigation, resources, data curation, writing – original draft, writing – review & editing, visualization, supervision, project administration

Kaitlin E Hanss: methodology, software, data curation, writing – review & editing Andrew J M Halls: methodology, writing – review & editing, supervision

Daniel F Becker: methodology, writing – review & editing, supervision

Anne L Glowinski: methodology, writing – review & editing, supervision

Andrew Krystal: methodology, resources, writing – review & editing, supervision, funding acquisition

## Abbreviations

AI: Artificial Intelligence
API: Application Programming Interface
DSM: Diagnostic and Statistical Manual
DSM-5-TR: Diagnostic and Statistical Manual, Version 5, Text Revision
GLMM: Generalized Linear Mixed Model
JSON: JavaScript Object Notation
LLM: Large Language Model
LRM: Large Reasoning Model
PPV: Positive Predictive Value
SR: Simulated Reasoning
SV: Self-Verification

## Multimedia Appendices

### Multimedia Appendix 1. Methods for diagnosis simplification and matching

#### Multimedia Appendix 2. Detailed scoring charts

This spreadsheet contains one tab with detailed scoring information for each combination of model and inference approach. The columns are as follows:

case_number: Vignette number from the DSM-5-TR Clinical Cases handbook. answers_dx: JSON string array containing the author-designated diagnoses model_dx: JSON string array containing the model predicted diagnoses

dx_TP: Count of true positive diagnoses

dx_TP_list: JSON string array containing the true positive diagnoses

dx_FP: Count of false positive diagnoses

dx_FP_list: JSON string array containing the false positive diagnoses

dx_FN: Count of false negative diagnoses

dx_FN_list: JSON string array containing the false negative diagnoses

dx_recall: The computed recall metric for this vignette

dx_precision: The computed precision metric for this vignette

