## Supplementary material for "Simulated Reasoning and Self-Verification for Psychiatric Diagnosis in Generalist Large Language Models: Comparative Evaluation": Multimedia Appendix 1

Simplification:

1. All DSM specifiers, modifiers and codes were removed
2. All neurocognitive disorders were collapsed into “Delirium” or “Neurocognitive Disorder,” removing disease-specific language and combining the mild and major classes.
3. If any diagnoses were identical after simplification, they were combined.

Matching:

First, all candidate diagnosis strings were scored for matching against the list of DSM-5-TR diagnoses using a fuzzy string matching algorithm (WRatio, rapidfuzz Python library). If the diagnosis exactly matched a single DSM-5-TR diagnosis (with strength >99%), that diagnosis was used. Otherwise, the diagnosis was added to a reconciliation list with the highest matching diagnosis and the matching score with that diagnosis.

The reconciliation list was then reviewed and reconciliations were resolved using the following process:

1. If the diagnosis was clearly non-psychiatric (e.g., “Hypertension”), the diagnosis was retained (as a false positive)
2. If the diagnosis was clearly related to a Z code (e.g., “Nonsuicidal Self-Injury”), the matching Z code diagnosis was used
3. If the diagnosis was clearly associated to one specific DSM-5-TR diagnosis but contained a minor typographical error or minor additional language (i.e., “Generalized Anxiety Disorder (GAD)” rather than “Generalized Anxiety Disorder), the diagnosis was normalized to the base diagnosis
4. If the diagnosis contained excess specifying language (i.e., “Major Depressive Disorder, severe” rather than “Major Depressive Disorder”) or was a specific instance of a diagnosis (i.e., “Cannabis-Induced Psychotic Disorder” rather than “Substance/Medication-Induced Psychotic Disorder), the diagnosis was normalized to the base diagnosis
5. If the diagnosis was clearly related to a diagnostic category but contained insufficient information to assign a specific diagnosis (i.e., “Depressive Episode”), the appropriate other/unspecified diagnosis and associated category were assigned (i.e., “Unspecified Depressive Disorder”)
6. If the diagnosis was a valid entity in a prior edition of the DSM that had been changed in the DSM-5-TR or clearly associated to an entity in the DSM-5-TR, the diagnosis was replaced with the associated new DSM-5-TR diagnosis and category
7. Otherwise, the diagnosis was retained (as a false positive) and assigned the category “Non-DSM”

All reconciliations were tracked with a numeric code, and once a specific reconciliation was established, it was put into a database and reused if the same string appeared again in the analysis for consistency.
